# Nutritional status of under-5 Rohingya children admitted for diarrheal diseases in primary health centers in Cox’s Bazar, Bangladesh

**DOI:** 10.1101/2023.06.05.23290974

**Authors:** Md. Fuad Al Fidah, Syeda Sumaiya Efa, Md. Emam Hossain, Tanvir Haider, Dipankor Roy

## Abstract

**Background:** One of the leading causes of mortality and morbidity in under-5 children is Diarrhea. Malnutrition, in association with the diarrheal disease, affects children negatively. In 2018, Bangladesh saw a massive influx of Rohingya people, with almost 29% of under-5 children.

**Objectives:** This study aimed to assess the nutritional status of under-5 Rohingya children admitted with acute diarrheal diseases at the primary health centers at Cox’s Bazar.

**Methods:** This cross-sectional study was conducted among the diarrhea affected under-5 Rohingya children who reported to primary health centers at cox’s bazar. The study was conducted from January to December 2018. The primary caregivers of 276 children who were affected with diarrhea were the respondents. Data were collected by face-to-face interview and record review using a semi-structured questionnaire and checklist respectively. All ethical issues were maintained strictly.

**Results:** The majority (60.1%) of the participants were male. Only 2.9% of the participants reported passing of visible blood in stool. Oral Rehydration Solution (ORS) was not taken by most of the participants (50.7%) before visit to the hospital. The mean (±SD) age was 18.54 (±12.44) months. Among the participants, 41.3% had underweight (<-2 WAZ score). Statistically, a significant relationship was found between the age of the participants and underweight (p<0.05) with a moderate effect size (Φ=0.21).

**Conclusion:** The study findings would be helpful to identify the need for raising awareness among caregivers of under-5 children regarding the use for ORS and contribute to reducing morbidity and mortality associated with malnutrition and diarrhea, and improving their overall health outcomes.

## Introduction

Diarrhea is a leading cause of death and morbidity in children under five years. Over 1,300 young infants die each day; that’s 526,000 a year, despite available basic and efficient treatment.^1^ Improved hygiene and water supply have reduced diarrheal mortality in recent decades. Unfortunately, diarrhea morbidity is still high.^2^ Bangladesh has an under-5 mortality rate of 35/1000 live births.^3^ 36.2% of under-5 children were stunted, 15% wasted, 33% underweight.^4^

Diarrhea and malnutrition are closely intertwined, forming a vicious cycle that can negatively impact the physical and cognitive development of young children.^5,6^ Diarrhea can cause nutrient deficiencies, leading to malnutrition. Conversely, as malnutrition impairs the immune system, children are more susceptible to diarrhea.^7^ It has long been understood that malnutrition and diarrheal illnesses are inversely correlated, with diarrhea being associated with reduced child growth, development, and cognitive performance.^8^ Recurrent episodes of diarrhea can particularly affect weight and height gain, further worsening malnutrition.^9^ Physical indicators of malnutrition like edema or wasting, can only be used to a certain extent, which emphasizes the necessity for precise anthropometric measurements to identify at-risk children.

Since the 1990s, many Rohingya have fled Myanmar to Bangladesh. During the 2017 Rakhine State violence, Rohingya refugees, including women and children, arrived in the country. The lack of basic facilities and poor living conditions in camps and settlements has increased health needs among these vulnerable refugees, who are often housed in designated sites or host communities. Recent rains and limited manpower, logistics, and finances make access to new settlements difficult. Thus, there are gaps in overall provision of life saving health care to affected populations and adjacent communities.

Various indicators are used to identify malnourished children under five years. Weight-for-age (W/A) is a child’s weight in relation to their age. This indicator shows whether a child is ‘moderately or severely underweight’. Assuming the child’s age is accurate, weight-for-age is a good predictor of acute malnutrition.^11^ World Health Organization’s (WHO) ‘WHO Anthro’ program (v3.2.2) can reliably measure weight-for-age z-score (WAZ) based on a patient’s weight and age. Weight fluctuates with time, reflecting acute malnutrition. Therefore, this study aimed to assess the nutritional status of under-5 children admitted for diarrheal diseases in primary health centers in Cox’s Bazar.

## Materials and Methods

### Study settings and subjects

We conducted this cross-sectional study for the period of one year, from January to December 2018. The study was conducted in two Rohingya camp areas: Balukhali and Samlapur. The study population was all ‘discharged’ under-5 Rohingya children recovered from diarrheal episode and treated at primary health centers in Cox’s Bazar. Caregivers of these children were the respondents.

### Sample size and sampling technique

The sample size of the study was 276. Convenience sampling technique was used.

### Data collection method and instruments

We used a semi-structured questionnaire and a checklist to collect data through face-to-face interview and reviewing available hospital records. Data was collected by the authors. WAZ score was calculated via ‘WHO Anthro’ software (v 3.2.2).

### Measurement

*Diarrheal Episode:* Three (3) or more loose stools lasting more than a day and separated from another episode by at least 48 hours or more without diarrhea.

### underweight

Children whose WAZ is below minus 2 (−2) standard deviations (SD) below the mean on the WHO Child Growth Standards.^11^

### Data management

Data were checked and verified at both field and central levels to ensure quality. Data were kept safely under the control of the principal investigator. We checked all data thoroughly to verify its relevancy and consistency. Incomplete and missing data were sorted out and verified. Data were coded, categorized, cleaned, and entered into IBM SPSS software (V 25). We carried out double data entry to perform a quality control check of the data.

### Statistical methods

The study used Statistical Package for Social Science (SPSS) software for data analysis. In the case of descriptive statistics, we estimated frequency distribution, percentage, mean, and standard deviation. Continuous data were reported in terms of mean and standard deviation, while categorical variables were portrayed by counts and percentages. In the case of inferential statistics, Chi-square test was used to find any association between categorical variables. A p-value <0.05 was considered statistically significant. All statistical tests were two-sided and performed at a significance level of α=0.05.

### Ethical considerations

Before commencing the study, the research protocol was approved by the research committee (Local ethical committee) of Rajshahi Medical College. Then it was assured that obtained information and records would be kept confidential, and would only be used for research purposes and the findings would be helpful for planning the management procedure of Rohingya children who are suffering from both diarrhea and malnutrition simultaneously.

## Results

Out of 276 participants, the majority (47.5%) was within the age group ≤12 months and 19.2% was in the age group of >24 months. mean (±SD) age was 18.54 (±12.44) months. Two-thirds (60.1%) of the participants were male. Most of the participants (68.1%) suffered from diarrhea for a total 12-72 hours with mean (±SD) duration of

39.87 hours (±37.61). Regarding underweight, most of the participants (58.7%) did not suffer from underweight. The percentage of underweight was 41.3%. The majority (97.1%) of the participants was suffering from acute watery diarrhea (AWD) and the rest had Invasive diarrhea. Most of the participants (50.7%) didn’t take ORS before admission (Table 1).

**Table 1:** Distribution of participants by background characteristics (n=276)

| Background characteristics | Category | f (%) |
| --- | --- | --- |
| Age group (Months) | $\leq 12$ | 131 (47.5) |
|  | 13-24 | 92 (33.3) |
| | $>24$ | 53 (19.2) |
| | Mean (±SD) | 18.54 (±12.44) |
| Sex | Male | 166 (60.1) |
|  | Female | 110 (39.9) |
| Duration of diarrhea (Hours) | $<12$ hours | 58 (21.3) |
|  | 12-72 hours | 188 (68.1) |
| | $>72$ hours | 30 (10.9) |
| | Mean (±SD) | 39.87 (±37.61) |
| Underweight | No | 162 (58.7) |
|  | Yes | 114 (41.3) |
| Type of diarrhea | Acute Watery diarrhea | 268 (97.1) |
|  | Invasive diarrhea | 8 (2.9) |
| ORS taken before coming to the hospital | Yes | 136 (49.3) |
|  | No | 140 (50.7) |

f: frequency; %: Percentage; SD: Standard deviation

By the age of the participants, underweight was significantly (p<0.05) higher in 13-24 months (37.7%). The effect size for this finding, Cramer’s V, was moderate (Φ=0.21, df=2). Age group ≤12 months were found to be second (36.0%). However, no significant association was found between sex, duration of diarrhea, type of diarrhea, ORS taken before coming to hospital and presence of malnutrition (Table 2).

**Table 2:**
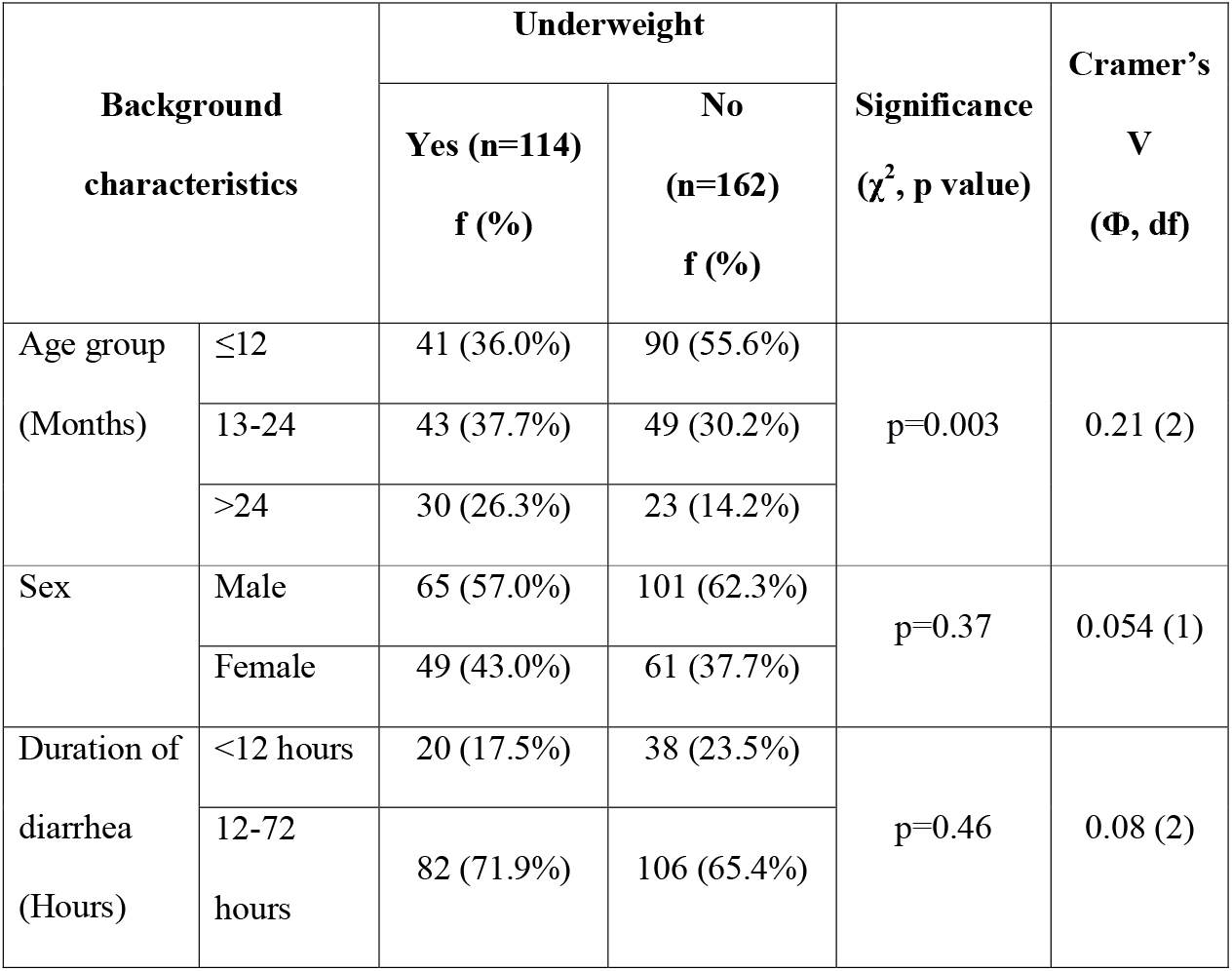
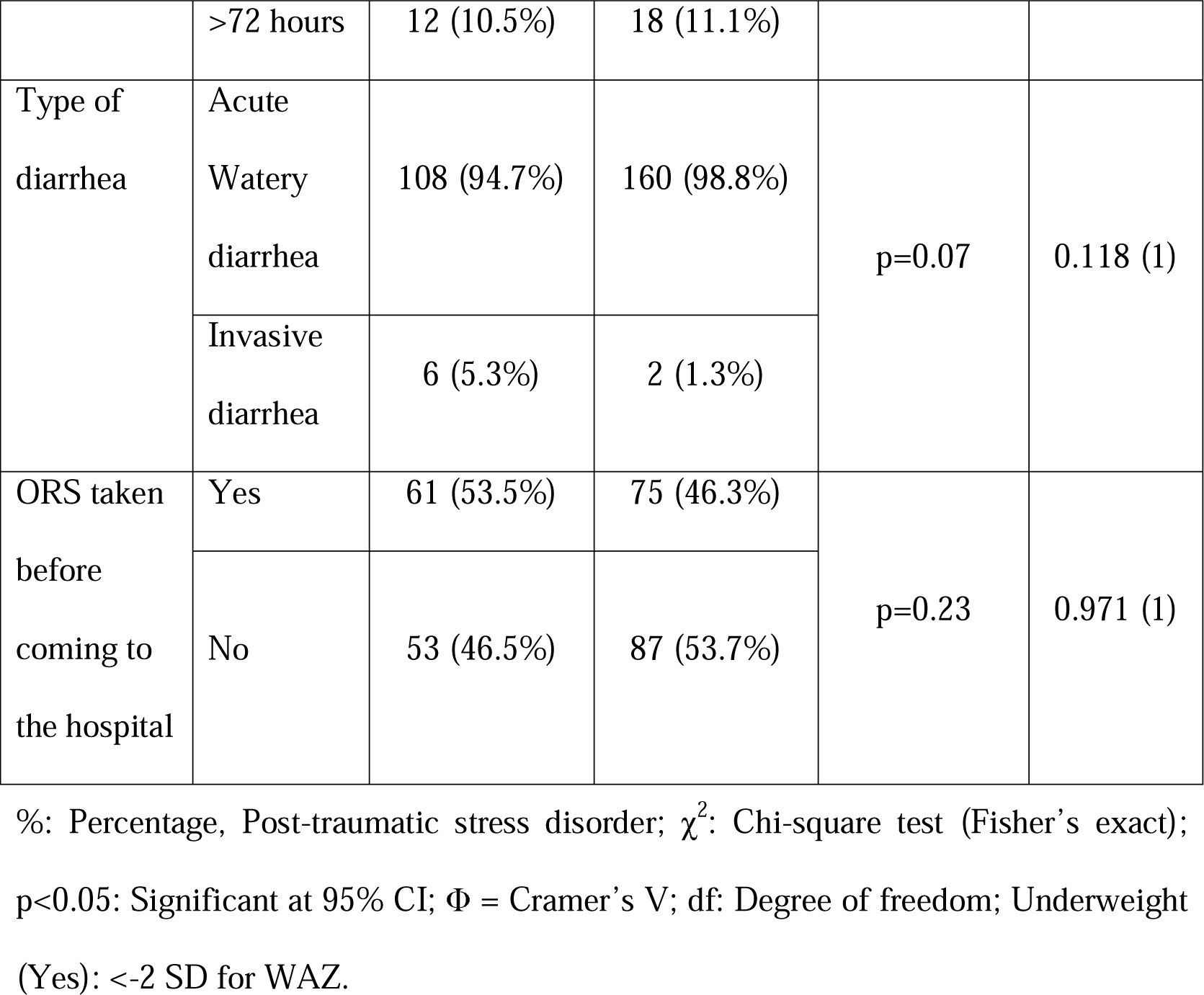
Association between underweight and selected background characteristics.

Univariate analysis was performed using the binary logistic regression model to determine the Odds Ratio (OR) for participants with underweight. Age group was incorporated into the model for underweight. The univariate analysis revealed that, children who belonged to the age group of 13-24 months and >24 months had higher odds of having underweight than children who aged ≤ 12 months. The OR was 1.93 (p-value: 0.02; 95% CI: 1.11-3.34) and 2.86 (p-value: 0.002; 95% CI: 1.49-5.52) respectively (Table 3).

**Table 3:**
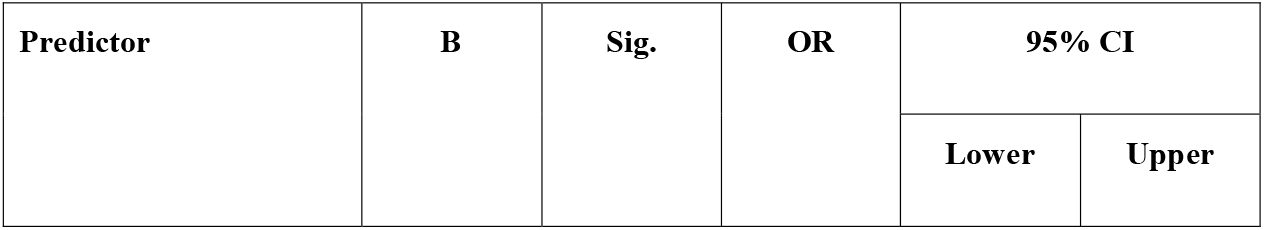
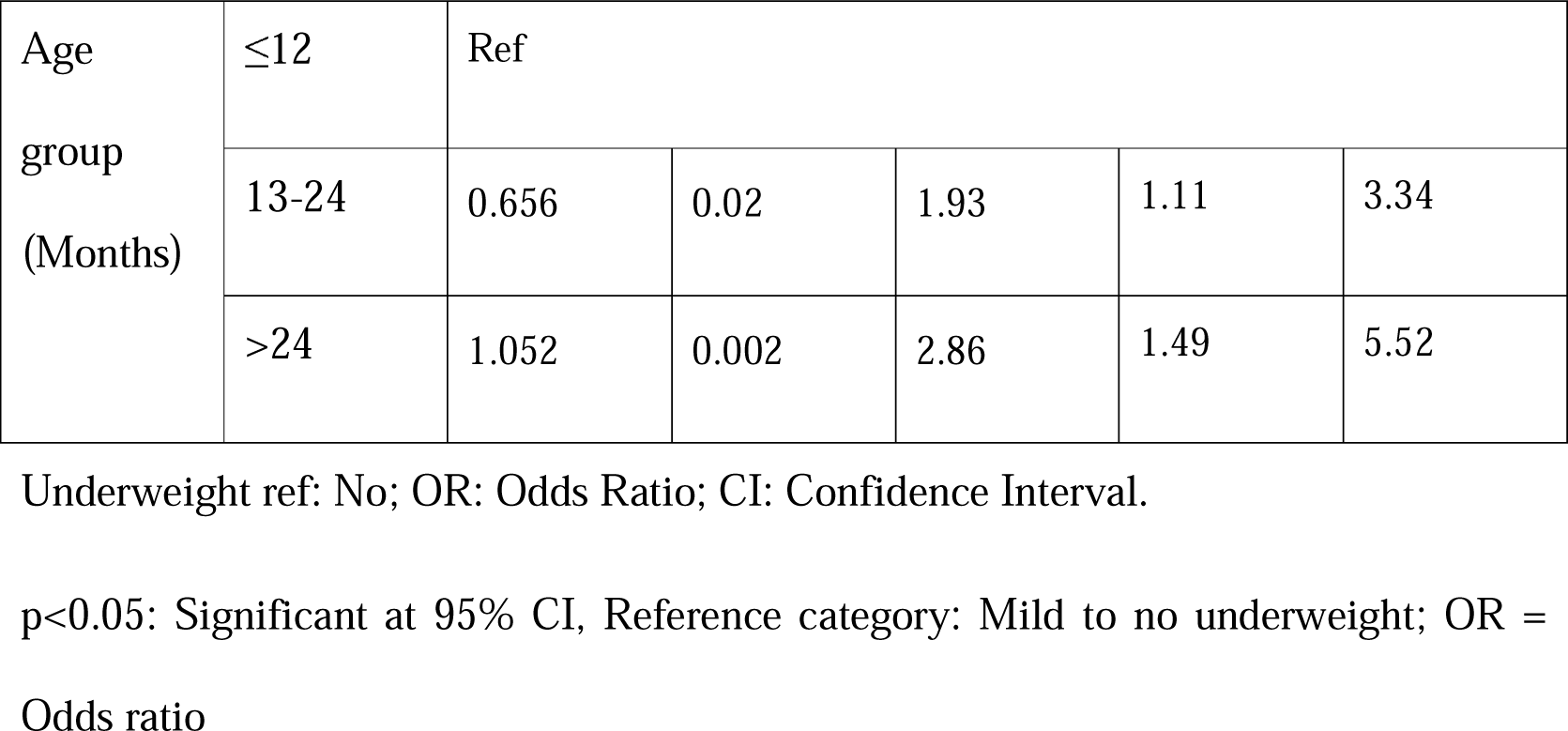
Binary logistic regression analysis of age associated Underweight.

| Predictor | B | Sig. | OR | 95% CI |  |
| --- | --- | --- | --- | --- | --- |
|  |  |  |  | Lower | Upper |

|  |  |  |  |  |  |  |
| --- | --- | --- | --- | --- | --- | --- |
| Age<br>group<br>(Months) | ≤12 | Ref |  |  |  |  |
|  | 13-24 | 0.656 | 0.02 | 1.93 | 1.11 | 3.34 |
|  | >24 | 1.052 | 0.002 | 2.86 | 1.49 | 5.52 |

Underweight ref: No; OR: Odds Ratio; CI: Confidence Interval.
p<0.05: Significant at 95% CI, Reference category: Mild to no underweight; OR = Odds ratio

## Discussion

This cross-sectional study was carried out to determine the nutritional status of under-5 Rohingya children who were admitted with diarrhea in primary health centers in Cox’s Bazar. The sample size was 276, participants were selected using convenience sampling.

Childhood malnutrition remains a significant public health concern in Bangladesh, as the prevalence of malnutrition remains high. A study conducted in Mirzapur, Bangladesh by Ferdous F et al. revealed that malnourished children, particularly those from resource-poor families, are at an increased risk of experiencing multiple episodes of diarrhea.^13^ The burden of diarrheal disease is particularly higher in settings with limited resources, such as emergency camps, where overcrowding is common. In such conditions, the Weight-for-Age (WAZ) indicator is commonly used to assess the relative body mass of children in relation to their age, as it serves as a useful tool to monitor growth and changes in the severity of malnutrition over time.

In the present study, out of 276 participants, the majority (46.7%) were in the age group of ≤12 months, which corresponds with Afroza Khatun’s study in Kushtia.^14^ The mean age of the participants was 18.54 (± 12.44 SD) months. The study also indicates that there is a preponderance of males (60.1%) over females (39.9%).

WHO defines diarrhea as the passing of three or more loose stools (which take the shape of the container) within 24 hours. A new episode of diarrhea can occur after two full days without diarrhea. Bouts of diarrhea lasting for less than 14 days are defined as acute; episodes continuing for more than 14 days are identified as persistent.^15^ In this study, we found that 97.1% of the participants were suffering from acute watery diarrhea, and only 2.9% of the participants were suffering from invasive diarrhea. Persistent diarrhea was not found in our study. However, a study conducted on the forcible displaced Myanmar nationals (FDMNs) reported that the prevalence of invasive diarrhea was 10.3% which does not correspond to our study. Possible explanation might be that our study was conducted on children under the age of 5, whereas age was not restricted in that study.^16^

We also found that 41.3% of children with diarrhea were suffering from underweight, which corresponds with the FDMN study that reports 40.9% presence of underweight.^16^ Another study conducted in South Africa reported that 35% of hospitalized children had moderate malnutrition, which supports the findings of our current study.^17^ Again, it has been shown that a child’s hydration state at the time of admission may have an effect on his or her nutritional status; therefore, this may have contributed to the high proportion since we only used admission weights.^18^ Most of the participants (68.1%) in our study had a history of duration of diarrhea for 13-72 hours.

Underweight was found to be statistically significant in relation to the age group of the participants (p <0.001). A higher age indicated higher chance of having underweight.

We can prevent most of the diarrheal deaths simply by prevention and treatment of dehydration. A study by Munos MK in 2010 stated that almost 93% of all diarrheal deaths might be prevented by using ORS.^19^ However, in this study, we have found that 50.7% of participants did not take ORS before coming to the hospital. This indicates a lack of knowledge regarding the use and effect of ORS among the caregivers.

The study had several limitations. As it required recall on the part of the participants, and their answers might have been influenced by recall bias. As the study was done in an emergency, there was a restriction on the movement of the Rohingya people. As a result, data regarding hospital stay may have been affected and thus excluded from the study. Also, discharge weights could have presented a clearer picture regarding prevalence of underweight as rehydration marked increases the weight of the diarrhea affected children who reported with some or severe dehydration.

## Conclusion

Malnutrition is a public health concern in Bangladesh. As the Rohingya people are forced to vacate their houses and take refugees in various camps in Bangladesh, various health-related issues impact their health. Diarrhea and malnutrition are two of them. Present study has demonstrated the prevalence of malnutrition among under-5 Rohingya children with diarrhea reporting to primary health centers in Cox’s bazar to be 41.3%. Age was found to be significantly associated with malnutrition, which highlights the need for age-specific interventions. It was also found that the caregivers of under-5 Rohingya children are reluctant or lack knowledge about ORS and the importance of its use during diarrhea. However, they do tend to seek health care for their children. The study suggests devising new policies to increase awareness regarding ORS use among Rohingya caregivers, and also proposes that early detection and prompt management of malnutrition should be integrated into the management of acute diarrhea in primary health centers. However, that study fails to consider weight during discharge. As hydration status is known to influence the weight of the patient, discharge weight would have been a better indicator to identify malnutrition.

## Data Availability

All data produced in the present study are available upon reasonable request to the authors

## Acknowledgement

The authors are indebted to all study participants and their caregivers for their participation. An earlier version of the current manuscript can be found as a preprint.^20^

